# The psychological effects of COVID-19 on frontline healthcare workers and how they are coping: a web-based, cross-sectional study from Pakistan

**DOI:** 10.1101/2020.06.03.20119867

**Authors:** Muhammad Salman, Muhammad Husnnain Raza, Zia Ul Mustafa, Tahir Mehmood Khan, Noman Asif, Humera Tahir, Naureen Shehzadi, Khalid Hussain

**Author notes:** **Corresponding author**. Address: Department of Pharmacy Practice, Faculty of Pharmacy, The University of Lahore, 1-Km Defence Road, Lahore, Pakistan. Email address.

## Abstract

**Background:** High level stress is expected when crises starts affecting people’s lives and communities which is witnessed in the past epidemics. Infectious diseases outbreaks like the ongoing COVID19 pandemic have negative impact on healthcare workers’ (HCWs) mental health, which needs to be investigated. Therefore, we aimed to assess the psychological impact of COVID-19 on frontline HCWs and their coping strategies.

**Methods:** A web-based, cross-sectional study was conducted among HCWs of the Punjab province of Pakistan. The generalized anxiety scale (GAD-7), patient health questionnaire (PHQ-9) and Brief-COPE were used to assess anxiety, depression and coping strategies.

**Results:** The mean age of respondents (N = 398) was 28.67 ± 4.15 years, with majority of medical doctors (52%). The prevalence of anxiety and depression were 21.4% and 21.9%, respectively. There was no significant difference of anxiety and depression scores among doctors, nurses and pharmacists. Females had significantly higher anxiety (p = 0.003) and depression (p = 0.001) scores than males. Moreover, HCWs performing duties in COVID-19 ICU had significantly higher anxiety score than those from isolation wards (p = 0.020) and other departments (p = 0.014). Depression, not anxiety, score were higher among those who did not receive the infection prevention training. Most frequently adopted coping strategy was religious coping (5.98 ± 1.73) followed by acceptance (5.59 ± 1.55) and coping planning (4.91 ± 1.85).

**Conclusion:** A considerable proportion of HCWs are having generalized anxiety and depression during the ongoing COVID-19 pandemic. Our findings call for interventions to mitigate mental health risks in HCWs.

## Introduction

All the measures taken to contain coronavirus disease (CVOID-19) since its emergence from Wuhan, in late December 2019, has not come to fruition yet. There were more than 4.8 million COVID-19 infected cases (Africa = 68347, Americas = 2166003, Eastern Mediterranean = 376379, Europe = 1946610, South-East Asia = 164225 and Western Pacific = 170910 cases) with 323256 deaths across the world as of May 21, 2020 [1]. In Pakistan, COVID-19 cases were estimated to be 50294 (active cases = 34,426, recoveries = 15,201, deaths = 1,067) as of May 22, 2020 [2].

Fighting at the forefronts, providing medical care to patients, healthcare workers (HCWs) are under constant threat of contracting the disease. Data from Italy, China, United States, Spain, and France showed that HCWs were increasingly being infected with COVID-19, ranging from 15-18% and in some cases up to 20% of the infected population [3]. In Pakistan, 766 HCWs have been infected with COVID-19, of them, 11 lost their life to it [4]. The continuous spread of COVID-19, risk of getting infected and transmitting it to love-ones, increased work load, physical exhaustion, shortage of personal protective equipment and the need to make ethically difficult decisions on rationing of care can profoundly influence the physical as well as mental well-being of HCWs [3, 5, 6]. This constant stress can trigger a variety of psychological issues such as anxiety, depression, sleep disturbances, panic attacks, posttraumatic stress symptoms, and avoidance of contact, depressive tendencies and helplessness [5]. The main objective of the current study was to assess the anxiety and depression among Pakistani HCWs during the ongoing COVID-19 pandemic. Secondary objective was the determination of coping strategies adopted by the HCWs to deal with it.

## Methods

A web-based, cross-sectional study was carried out among HCWs of the Punjab province of Pakistan from April 15-May 20, 2020. In the present study, the medical doctors, nurses and pharmacist were eligible for inclusion. Excluded were those who were unwilling to participate, could not understand English language, healthcare assistants and other hospital staff. This study was approved by the Research Ethics Committee of the Department of Pharmacy Practice, Faculty of Pharmacy, The University of Lahore. An online informed consent was obtained from every participant prior to their enrolment.

An online self-administered questionnaire was deigned to assess the psychological effects of COVID-19 on HCWs and coping strategies adopted by them. As the medical, pharmacy and nursing education in Pakistan are entirely English-medium, there was no need to translate the questionnaire into Urdu language. The content of questionnaire was validated by an expert panel. Moreover, the questionnaire was pilot tested among ten HCWs (5 medical doctors, 2 nurses and 3 pharmacists) who reported no problems in understanding questions as well as their response categories. Google forms were used to disseminate the online questionnaire.

We used generalized anxiety scale (GAD-7) to assess anxiety among respondents [7]. It had seven items, each of which was scored 0 (not at all) to 3 (nearly every day), providing a 0 to 21 score. The total score was categorized into four severity groups: minimal-none (≤ 4), mild (5-9), moderate (10-14) and severe (≥ 15). The Patient health questionnaire (PHQ-9) was used to assess depression. It contained 9-items, each of which was scored 0 (not at all) to 3 (nearly every day), providing a 0-27 score. The total score was categorized into five severity groups: minimal-none (≤ 4), mild (5-9), moderate (10-14), moderately severe (15-19) and severe (≥ 20) [8]. In the present study, respondents’ achieving score ≥ 10 on GAD-7 and PHQ-9 were considered as having anxiety and depression, respectively.

The Brief-COPE scale was used to assess the coping strategies adopted by HCWs during COVID-19 pandemic [9]. It contained 28-items, each of which was scored from 1 (I have not been doing this at all) to 4 (I have been doing this a lot). This scale explored the following 14 coping methods: self-distraction, active coping, denial, substance use, use of emotional support, use of instrumental support, behavioural disengagement, venting, positive reframing, planning, humour, acceptance, religion and self-blame. Possible scores for each subscale ranged from 2 to 8, with higher scores indicating a higher tendency to implement the corresponding coping style.

All the data were entered and analysed using IBM SPSS version 22 for Windows. Continuous data were presented as mean and standard deviation (SD) whereas categorical data were expressed as number and percentages. A p-value of less than 0.05 was considered statistically significant. Independent t-test and ANOVA test were performed, where applicable, to compare difference of anxiety, depression and coping strategies scores among demographic variables. Moreover, for tri- or polychotomus variables, a series of post-hoc analysis with Bonferroni adjustment were performed to assess significance among intergroup variables.

## Results

A total of 398 responses were included in the analysis. Demographics of the respondents are shown in Table 1. Mean age of respondents was 28.67 ± 4.15 years, with majority of females (56%). Around 52% were medical doctors whereas nurses and pharmacists were 33.4% and 15.1%, respectively. Approximately 28% respondents were performing duties in COVID-19 isolation wards and 5% were providing medical care in CVOID-19 intensive care units.

**Table 1:** Anxiety and depression assessments based on respondents’ demographics.

| Variable | N (%) | Anxiety score<br>Mean ± SD | Depression score<br>Mean ± SD |
| --- | --- | --- | --- |
| **Age (years)** |  |  |  |
| ≤ 25 | 81 (20.4) | 6.57 ± 4.21 | 7.35 ± 4.95 |
| 26-30 | 213 (53.5) | 6.74 ± 4.41 | 6.59 ± 5.31 |
| 31-35 | 84 (21.1) | 7.79 ± 4.89 | 7.00 ± 5.21 |
| >35 | 20 (5.0) | 4.75 ± 2.81* | 5.25 ± 3.29 |
| **Gender** |  |  |  |
| Male | 183 (46.0) | 6.10 ± 4.281 | 5.81 ± 5.13 |
| Female | 215 (54.0) | 7.44 ± 4.498** | 7.48 ± 5.02† |
| **Occupation** |  |  |  |
| Doctor | 205 (51.5) | 6.52 ± 4.38 | 6.60 ± 5.45 |
| Nurse | 133 (33.4) | 7.23 ± 4.27 | 7.22 ± 4.06 |
| Pharmacist | 60 (15.1) | 6.98 ± 5.01 | 6.00 ± 6.03 |
| **Experience (years)** |  |  |  |
| ≤ 5 | 302 (75.9) | 6.67 ± 4.37 | 6.62 ± 5.19 |
| 6-10 | 77 (19.3) | 7.44 ± 4.75 | 7.39 ± 5.20 |
| > 10 | 19 (4.8) | 6.89 ± 4.37 | 5.42 ± 3.42 |
| **Placement** |  |  |  |
| Isolation ward/quarantine | 113 (28.4) | 6.68 ± 4.84 | 7.67 ± 6.20 |
| COVID-19 intensive care unit | 20 (5.0) | 9.60 ± 4.08* | 8.10 ± 1.04* |
| Other areas | 265 (66.6) | 6.68 ± 4.23 | 6.20 ± 0.28 |
| **Infection prevention training**<br>**for COVID-19** |  |  |  |
| Yes | 131 (32.9) | 6.61 ± 4.26 | 5.71 ± 4.58 |
| No | 267 (67.1) | 6.93 ± 4.54 | 7.21 ± 2.33** |

\*P-value < 0.05; \*\*P-value < 0.01; †P-value ≤ 0.001; One-way ANOVA was applied for three or more groups. To estimate the difference among two groups independent sample t-test was applied. P-value <0.05 was considered statistically significant.

The mean anxiety and depression score were 6.83 ± 4.44 and 6.72 ± 5.14, respectively. The frequency of respondents’ having none-mild, moderate, and severe anxiety were 78.7%, 13.1% and 8.3%, respectively. Regarding the severity of depression, 21.8% scored equal or above 10 on PHQ-9 (minimal-none 35.9%, mild 42.2%, moderate 12.8%, moderately severe 7.3%, and severe 1.8%). Comparison of anxiety and depression scores among respondents’ demographics are shown in Table 1. Female respondents were found to have significantly higher anxiety (p = 0.003) and depression (p = 0.001) scores than male respondents. Significant differences of anxiety score were observed in age as well as placement categories (p = 0.016). In post-hoc analysis with Bonferroni correction (Table 2), HCWs above 35 years of age had significantly lower anxiety score than those belonging to 31-35 years age group. Moreover, HCWs providing services in ICU had significantly higher anxiety score than those from isolation wards (p = 0.020) and other departments (p = 0.014). As shown in Table 3, around 79% respondents reported fear of contracting the infection and 81.9% were afraid their love-ones might get infected because of them. Moreover, shortage of protective equipment and lack of mass-scale COVID-19 screening in Pakistan were major distresses.

**Table 2:** Post-hoc analysis with Bonferroni correction for anxiety and depression scores among age and placement categories.

| Multiple-comparisons | Anxiety score | Depression score |
| --- | --- | --- |
|  | P-value | P-value |
| **Age (years)** |  | -- |
| ≤ 25 vs 26-30 | 1.000 |  |
| ≤ 25 vs 31-35 | 0.463 |  |
| ≤ 25 vs > 35 | 0.598 |  |
| 26-30 vs 31-35 | 0.402 |  |
| 26-30 vs > 35 | 0.326 |  |
| 31-35 vs >35 | **0.036** |  |
| **Placement** |  |  |
| Isolation ward vs COVID-19 intensive care unit | **0.020** | 1.000 |
| Isolation ward vs Other areas | 1.000 | **0.032** |
| COVID-19 intensive care unit vs Other areas | 0.014 | 0.328 |

**Table 3:** Sources of distress among respondents.

|  | N (%) |
| --- | --- |
| Afraid of getting infected any moment | 314 (78.9) |
| Afraid of bringing the virus to home | 326 (81.9) |
| Poor relationship between family members/friends and me induced by COVID-19 | 211 (53.0) |
| Worried because public in general is careless and not complying with disease control measures | 317 (79.6) |
| Afraid the disease will persist in Pakistani community for a very long time | 304 (76.4) |
| Worried about lack of protective equipment | 324 (81.4) |
| Worried about large-scale diagnostic testing for COVID-19 is not being done in Pakistan | 332 (83.4) |
| Incapability managing critical COVID-19 patients | 224 (56.3) |

As shown in Table 4, mean score was highest for religious coping (5.98 ± 1.73) followed by acceptance (5.59 ± 1.55) and coping planning (4.91± 1.85) whereas it was the lowest for substance use (2.95 ± 1.06) followed by self-blame (2.90 ± 1.29). Between-demographic differences of various coping strategies are also shown in Table 4. There was no significant difference of coping styles among age categories except for substance use (p = 0.004). Females were observed to have significantly higher scores for behavioural disengagement (p = 0.043), venting (p = 0.015) and religious/spiritual coping (p = 0.003) than male respondents. Amongst HCWs categories, statistically significant difference were seen for self-blame (p = 0.029), denial (p = < 0.001), substance use (p = 0.001), seeking emotional support (p = 0.033) and behavioural disengagement (p = 0.028). Furthermore, there was no significant difference of all the coping styles among HCWs working in isolation wards of hospitals, COVID-19 ICU and other areas except for substance use (p = 0.031) and positive reframing (p = 0.041). As presented in Table 5, a series of post hoc test with Bonferroni adjustment were carried out to determine significance among intergroup variables. Respondents belonging to 26-30 years age group reported significantly less substance use than those from 31-35 years of age (p = 0.020). Lastly, nurses had significantly higher coping style scores on denial, substance use and behavioural disengagement than doctors, however, there was no difference between doctors and pharmacists as well as nurses and pharmacists (Table 5).

**Table 4:**
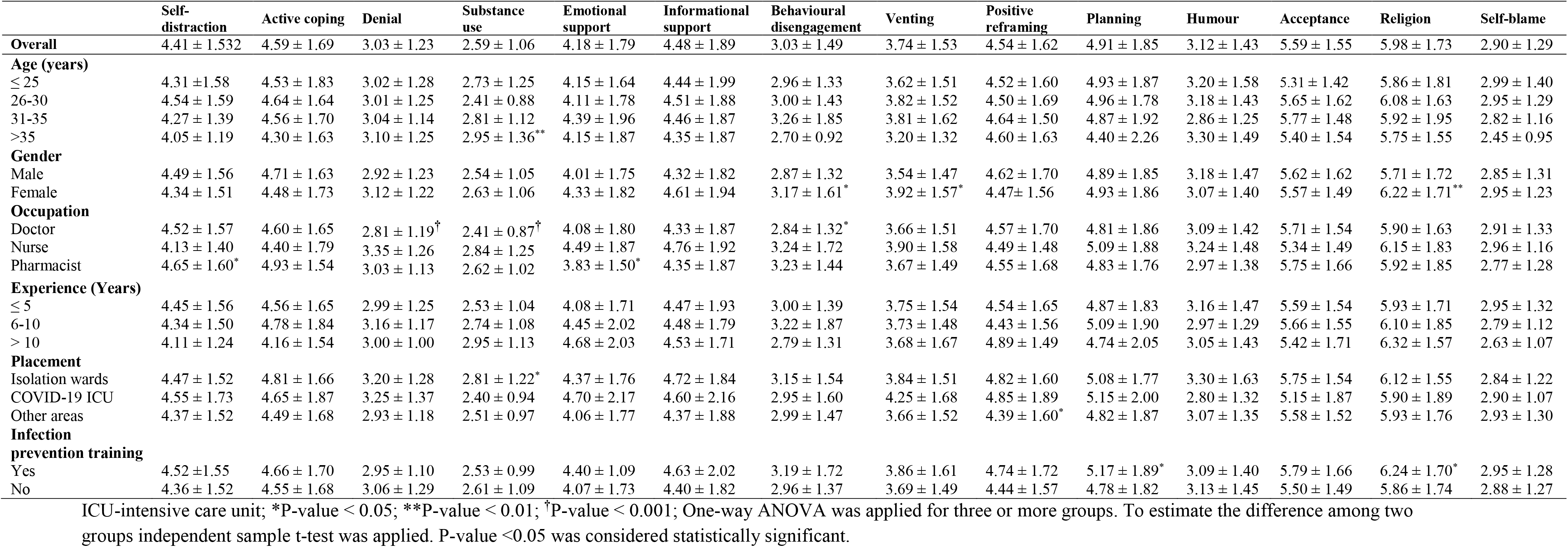
Coping Strategies adopted by the study participants.

|  | Self-distraction | Active coping | Denial | Substance use | Emotional support | Informational support | Behavioural disengagement | Venting | Positive reframing | Planning | Humour | Acceptance | Religion | Self-blame |
| --- | --- | --- | --- | --- | --- | --- | --- | --- | --- | --- | --- | --- | --- | --- |
| **Overall** | 4.41 ± 1.532 | 4.59 ± 1.69 | 3.03 ± 1.23 | 2.59 ± 1.06 | 4.18 ± 1.79 | 4.48 ± 1.89 | 3.03 ± 1.49 | 3.74 ± 1.53 | 4.54 ± 1.62 | 4.91 ± 1.85 | 3.12 ± 1.43 | 5.59 ± 1.55 | 5.98 ± 1.73 | 2.90 ± 1.29 |
| **Age (years)** |  |  |  |  |  |  |  |  |  |  |  |  |  |  |
| ≤ 25 | 4.31 ± 1.58 | 4.53 ± 1.83 | 3.02 ± 1.28 | 2.73 ± 1.25 | 4.15 ± 1.64 | 4.44 ± 1.99 | 2.96 ± 1.33 | 3.62 ± 1.51 | 4.52 ± 1.60 | 4.93 ± 1.87 | 3.20 ± 1.58 | 5.31 ± 1.42 | 5.86 ± 1.81 | 2.99 ± 1.40 |
| 26-30 | 4.54 ± 1.59 | 4.64 ± 1.64 | 3.01 ± 1.25 | 2.41 ± 0.88 | 4.11 ± 1.78 | 4.51 ± 1.88 | 3.00 ± 1.43 | 3.82 ± 1.52 | 4.50 ± 1.69 | 4.96 ± 1.78 | 3.18 ± 1.43 | 5.65 ± 1.62 | 6.08 ± 1.63 | 2.95 ± 1.29 |
| 31-35 | 4.27 ± 1.39 | 4.56 ± 1.70 | 3.04 ± 1.14 | 2.81 ± 1.12 | 4.39 ± 1.96 | 4.46 ± 1.87 | 3.26 ± 1.85 | 3.81 ± 1.62 | 4.64 ± 1.50 | 4.87 ± 1.92 | 2.86 ± 1.25 | 5.77 ± 1.48 | 5.92 ± 1.95 | 2.82 ± 1.16 |
| >35 | 4.05 ± 1.19 | 4.30 ± 1.63 | 3.10 ± 1.25 | 2.95 ± 1.36** | 4.15 ± 1.87 | 4.35 ± 1.87 | 2.70 ± 0.92 | 3.20 ± 1.32 | 4.60 ± 1.63 | 4.40 ± 2.26 | 3.30 ± 1.49 | 5.40 ± 1.54 | 5.75 ± 1.55 | 2.45 ± 0.95 |
| **Gender** |  |  |  |  |  |  |  |  |  |  |  |  |  |  |
| Male | 4.49 ± 1.56 | 4.71 ± 1.63 | 2.92 ± 1.23 | 2.54 ± 1.05 | 4.01 ± 1.75 | 4.32 ± 1.82 | 2.87 ± 1.32 | 3.54 ± 1.47 | 4.62 ± 1.70 | 4.89 ± 1.85 | 3.18 ± 1.47 | 5.62 ± 1.62 | 5.71 ± 1.72 | 2.85 ± 1.31 |
| Female | 4.34 ± 1.51 | 4.48 ± 1.73 | 3.12 ± 1.22 | 2.63 ± 1.06 | 4.33 ± 1.82 | 4.61 ± 1.94 | 3.17 ± 1.61* | 3.92 ± 1.57* | 4.47 ± 1.56 | 4.93 ± 1.86 | 3.07 ± 1.40 | 5.57 ± 1.49 | 6.22 ± 1.71** | 2.95 ± 1.23 |
| **Occupation** |  |  |  |  |  |  |  |  |  |  |  |  |  |  |
| Doctor | 4.52 ± 1.57 | 4.60 ± 1.65 | 2.81 ± 1.19<sup>†</sup> | 2.41 ± 0.87<sup>†</sup> | 4.08 ± 1.80 | 4.33 ± 1.87 | 2.84 ± 1.32* | 3.66 ± 1.51 | 4.57 ± 1.70 | 4.81 ± 1.86 | 3.09 ± 1.42 | 5.71 ± 1.54 | 5.90 ± 1.63 | 2.91 ± 1.33 |
| Nurse | 4.13 ± 1.40 | 4.40 ± 1.79 | 3.35 ± 1.26 | 2.84 ± 1.25 | 4.49 ± 1.87 | 4.76 ± 1.92 | 3.24 ± 1.72 | 3.90 ± 1.58 | 4.49 ± 1.48 | 5.09 ± 1.88 | 3.24 ± 1.48 | 5.34 ± 1.49 | 6.15 ± 1.83 | 2.96 ± 1.16 |
| Pharmacist | 4.65 ± 1.60* | 4.93 ± 1.54 | 3.03 ± 1.13 | 2.62 ± 1.02 | 3.83 ± 1.50* | 4.35 ± 1.87 | 3.23 ± 1.44 | 3.67 ± 1.49 | 4.55 ± 1.68 | 4.83 ± 1.76 | 2.97 ± 1.38 | 5.75 ± 1.66 | 5.92 ± 1.85 | 2.77 ± 1.28 |
| **Experience (Years)** |  |  |  |  |  |  |  |  |  |  |  |  |  |  |
| ≤ 5 | 4.45 ± 1.56 | 4.56 ± 1.65 | 2.99 ± 1.25 | 2.53 ± 1.04 | 4.08 ± 1.71 | 4.47 ± 1.93 | 3.00 ± 1.39 | 3.75 ± 1.54 | 4.54 ± 1.65 | 4.87 ± 1.83 | 3.16 ± 1.47 | 5.59 ± 1.54 | 5.93 ± 1.71 | 2.95 ± 1.32 |
| 6-10 | 4.34 ± 1.50 | 4.78 ± 1.84 | 3.16 ± 1.17 | 2.74 ± 1.08 | 4.45 ± 2.02 | 4.48 ± 1.79 | 3.22 ± 1.87 | 3.73 ± 1.48 | 4.43 ± 1.56 | 5.09 ± 1.90 | 2.97 ± 1.29 | 5.66 ± 1.55 | 6.10 ± 1.85 | 2.79 ± 1.12 |
| > 10 | 4.11 ± 1.24 | 4.16 ± 1.54 | 3.00 ± 1.00 | 2.95 ± 1.13 | 4.68 ± 2.03 | 4.53 ± 1.71 | 2.79 ± 1.31 | 3.68 ± 1.67 | 4.89 ± 1.49 | 4.74 ± 2.05 | 3.05 ± 1.43 | 5.42 ± 1.71 | 6.32 ± 1.57 | 2.63 ± 1.07 |
| **Placement** |  |  |  |  |  |  |  |  |  |  |  |  |  |  |
| Isolation wards | 4.47 ± 1.52 | 4.81 ± 1.66 | 3.20 ± 1.28 | 2.81 ± 1.22* | 4.37 ± 1.76 | 4.72 ± 1.84 | 3.15 ± 1.54 | 3.84 ± 1.51 | 4.82 ± 1.60 | 5.08 ± 1.77 | 3.30 ± 1.63 | 5.75 ± 1.54 | 6.12 ± 1.55 | 2.84 ± 1.22 |
| COVID-19 ICU | 4.55 ± 1.73 | 4.65 ± 1.87 | 3.25 ± 1.37 | 2.40 ± 0.94 | 4.70 ± 2.17 | 4.60 ± 2.16 | 2.95 ± 1.60 | 4.25 ± 1.68 | 4.85 ± 1.89 | 5.15 ± 2.00 | 2.80 ± 1.32 | 5.15 ± 1.87 | 5.90 ± 1.89 | 2.90 ± 1.07 |
| Other areas | 4.37 ± 1.52 | 4.49 ± 1.68 | 2.93 ± 1.18 | 2.51 ± 0.97 | 4.06 ± 1.77 | 4.37 ± 1.88 | 2.99 ± 1.47 | 3.66 ± 1.52 | 4.39 ± 1.60* | 4.82 ± 1.87 | 3.07 ± 1.35 | 5.58 ± 1.52 | 5.93 ± 1.76 | 2.93 ± 1.30 |
| **Infection prevention training** |  |  |  |  |  |  |  |  |  |  |  |  |  |  |
| Yes | 4.52 ± 1.55 | 4.66 ± 1.70 | 2.95 ± 1.10 | 2.53 ± 0.99 | 4.40 ± 1.09 | 4.63 ± 2.02 | 3.19 ± 1.72 | 3.86 ± 1.61 | 4.74 ± 1.72 | 5.17 ± 1.89* | 3.09 ± 1.40 | 5.79 ± 1.66 | 6.24 ± 1.70* | 2.95 ± 1.28 |
| No | 4.36 ± 1.52 | 4.55 ± 1.68 | 3.06 ± 1.29 | 2.61 ± 1.09 | 4.07 ± 1.73 | 4.40 ± 1.82 | 2.96 ± 1.37 | 3.69 ± 1.49 | 4.44 ± 1.57 | 4.78 ± 1.82 | 3.13 ± 1.45 | 5.50 ± 1.49 | 5.86 ± 1.74 | 2.88 ± 1.27 |

ICU-intensive care unit; \*P-value < 0.05; \*\*P-value < 0.01; <sup>†</sup>P-value < 0.001; One-way ANOVA was applied for three or more groups. To estimate the difference among two groups independent sample t-test was applied. P-value < 0.05 was considered statistically significant.

**Table 5:** Multiple comparisons of coping strategies among selected variables using Bonferroni correction.

| Multiple comparisons | Self-distraction | Denial | Substance<br>use | Emotional<br>support | Behavioural<br>disengagement | Positive<br>reframing |
| --- | --- | --- | --- | --- | --- | --- |
| **Age (years)** | -- | -- |  | -- | -- | -- |
| ≤ 25 vs 26-30 |  |  | 0.126 |  |  |  |
| ≤ 25 vs 21-35 |  |  | 1.000 |  |  |  |
| ≤ 25 vs > 35 |  |  | 1.000 |  |  |  |
| 26-30 vs 31-35 |  |  | **0.020** |  |  |  |
| 26-30 vs > 35 |  |  | 0.169 |  |  |  |
| 31-35 vs > 35 |  |  | 1.000 |  |  |  |
| **Occupation** |  |  |  |  |  | -- |
| Doctor vs Nurse | 0.062 | < **0.001** | **0.001** | 0.125 | **0.046** |  |
| Doctor vs Pharmacist | 1.000 | 0.653 | 0.560 | 1.000 | 0.212 |  |
| Nurse vs Pharmacist | 0.084 | 0.290 | 0.493 | 0.056 | 1.000 |  |
| **Placement** | -- | -- |  | -- | -- |  |
| Isolation ward vs COVID-19 intensive care unit |  |  | 0.336 |  |  | 1.000 |
| Isolation ward vs Other areas |  |  | **0.037** |  |  | 0.054 |
| COVID-19 intensive care unit vs Other areas |  |  | 1.000 |  |  | 0.668 |

## Discussion

A recent position paper has underscored the dire need of quality data on the psychological effects of the COVID-19 pandemic across the whole population and vulnerable groups, and on brain function, cognition, and mental health of patients with COVID-19 [10]. Since HCWs are amongst the high-risk groups for getting COVID-19 and are particularly vulnerable to a variety of mental health problems, the present study was undertaken to provide insight on the impact of COVID-19 pandemic on mental well-being of Pakistani doctors, nurses and pharmacists fighting at the forefronts. Our findings revealed that prevalence of anxiety and depression were 21.4% and 21.9%, respectively. A recent review article identified 12 studies (11 from China [11-20] and 1 from Singapore [21]) that estimated anxiety and 10 studies (9 among Chinese HCWs and 1 amongst Singaporean HCWs) that estimated depression among HCWs [11, 12, 14, 15, 17, 18-21]. The calculated pooled prevalence of anxiety and depression was 23.2% (95% CI 17.77-29.12) and 22.8% (95% CI 15.1-31.51), respectively. Regarding the impact of anxiety and depressive symptoms on the quality of life, 51.8%, 6.3% and 1.5% reported somewhat, very, and extreme difficulty in doing work, taking care of things at home, or getting along with others.

Similar to findings of recently published studies [18, 19], stressors related to COVID-19 in our respondents included fear of getting infected with the deadly virus and transmitting it to their family members, poor relationship between family/relatives/friends and them induced by COVID-19, worry about public not complying with disease control measures, fear that the disease will not be controlled and persist in the country for a very long time, shortage of protective equipment, and apprehension over large-scale COVID-19 diagnostic testing not being done. Seven hundred sixty six HCWs have been infected with COVID-19 in Pakistan, of them, 11 has lost their life [4]. Moreover, family members of the HCWs who have died were also tested positive for COVID-19 [22]. Pakistani doctors and nurses have been protesting due to the insufficient provision of personal protective equipment, going on hunger strike and also threatening to quit working [22-25]. Additionally, level of COVID-19 testing in Pakistan has been reported to be low as compared to other countries [26]. Our findings highlight the need to not only providing adequate psychological support to our frontline medical forces via print, electronic and social media but also take all the measures necessary to reduce other stressors.

Regarding the coping strategies, Eisenberg and colleagues have reported two major components namely “avoidant coping” and “approach coping” in the Brief-COPE [27]. The humour and religion subscales were not included in either component as they did not exclusively load on either of the abovementioned components. Avoidant Coping was described by the subscales of denial, substance use, venting, behavioural disengagement, self-distraction, and self-blame. These coping styles are not ideal at managing anxiety and stress [27]. On the other hand, approach coping is characterized by the subscales of active coping, positive reframing, planning, acceptance, seeking emotional, and informational support. Compared to avoidant coping, these have been associated with better responses to adversity, including adaptive practical adjustment, better physical health outcomes and more stable emotional responding. Meyer categorized the strategies measured by the Brief-COPE into maladaptive and adaptive coping [28]. In addition to other subscales, religion and humour were considered as adaptive coping. In the present study, it was encouraging to see that our respondents’ scores for positive coping strategies were greater than avoidant or maladaptive coping.

Although we achieved our study objectives, our study had several limitations. First, this study was conducted among HCWs of only the Punjab province of Pakistan so our findings may not be generalized to overall HCWs population of Pakistan. Second, the survey was administered using snowball sampling method and we used a self-administered questionnaire so disadvantages associated with self-report data (e.g. introspective ability, response bias and sampling bias) may be present. Third, the clinical assessment for the diagnosis of depression and generalized anxiety disorders as per criteria of Diagnostic and Statistical Manual of Mental Disorders was not done. However, our findings offer valuable insight about the psychological impact of COVID-19 on frontline medical forces, sources of distress, and coping strategies.

## Conclusions

Our study reveals that 21.4% and 21.9% healthcare professionals have generalized anxiety and depression during the COVID-19 pandemic, respectively. Most frequently adopted coping strategies are religious/spiritual coping, acceptance, planning, active coping and positive reframing. Findings draw attention to proactively take steps to protect the mental wellbeing, enhance resilience and mitigate vulnerability of healthcare forces during COVID-19.

## Data Availability

The data of the current study are available from the corresponding author on reasonable request.

## Acknowledgments

We are grateful to the respondents for sparing their precious time to take part in the study.

## Authors’ contributions

MS, TMK and MHR conceived and designed the study. MS, ZUM and NA conducted literature review and designed the study questionnaire. MS, NS, TMK and HT conducted statistical analyses, with additional advice regarding analyses contributed by KH. MS drafted the manuscript, and all authors contributed to editing it and approved the final manuscript.

